# Time-specific effects of a multifaceted intervention on accelerometer-measured physical activity in primary school children: A cluster-randomized controlled trial

**DOI:** 10.1101/2022.05.06.22274770

**Authors:** Zheng Liu, Zhi-Han Yue, Li-Ming Wen, Jinfeng Zhao, Shuang Zhou, Ai-Yu Gao, Fang Zhang, Hai-Jun Wang

## Abstract

**Background:** It is unclear whether intervention effects on school-aged children’s physical activity differ across specific periods of the week or day. This study aimed to assess the time-specific intervention effects on accelerometer-measured physical activity in primary school children.

**Methods:** This was a nested study in a cluster-randomized controlled trial conducted from September 2018 to June 2019 in China. The intervention group included 4 schools (119 children) and the control group included 4 schools (99 children) in Beijing. The obesity prevention intervention engaged schools and families to improve children’s physical activity. Outcome measures included accelerometer-assessed intensity and amounts of physical activity within specific periods of a week (weekday/weekend day) or a day (in-school/out-of-school periods). Linear mixed models were used to estimate intervention effects. Subgroup analyses were also conducted to examine potential moderators including sex, age, body mass index, physical activity, and accelerometer compliance.

**Results:** The intervention lead to an increase in time engaged in moderate-to-vigorous physical activity (MVPA) within in-school periods of a day (adjusted mean difference: 0.54 minutes/hour; 95% confidence interval: 0.13, 0.94, *P* = 0.012) but it did not improve physical activity within out-of-school periods (*P* > 0.05) compared with the control group. There was no difference in the effect size across most of the moderators except for age, as younger children appeared to benefit more from the intervention in the improvement of in-school MVPA (*P*_interaction_ = 0.035). No between-group differences were observed in physical activity within the whole weekday or weekend day (*P* > 0.05).

**Conclusion:** The intervention effectively increased MVPA within in-school periods but did not improve out-of-school physical activity. Findings support the tailoring of intervention components to specific periods of a day to improve school-aged children’s whole pattern of physical activity.

**TRIAL REGISTRATION:** ClinicalTrials.gov Identifier: NCT03665857

## Introduction

Globally, more than three-quarters (81%) of children and adolescents are insufficiently physically active [1]. Sedentary behavior in children is associated with harmful effects on adiposity, physical fitness, cardiometabolic health, social functioning, and sleep [2, 3]. In comparison, physical activity, especially of moderate-to-vigorous intensity, confers health benefits for bone health, cognitive outcomes, and mental health in addition to physical fitness and cardiometabolic health [3, 4]. Global action on physical activity has been approved by the World Health Organization (WHO) to reduce the prevalence of physical inactivity by 15% by 2030 [5].

Interventions are urgently needed to effectively improve physical activity in children. Findings from recent systematic reviews and meta-analyses show that interventions only have small to negligible effects on improving physical activity in children [6, 7]. Despite the limited success in changing the whole physical activity in children, observational evidence indicates that intervention might be more beneficial during specific periods of the week (i.e., weekday/weekend day) or the day (i.e., in-school/out-of-school periods)[8]. However, only a paucity of intervantions focusing on childhood obesity prevention have examined the time-specific effects on physical activity until now [9]. Among others, one of the critical barriers was the availability of a measurement tool that can accurately assess the real-time response to an intervention.

It is vital to capture the physical activity within specific periods of a week or day during the implementation of an intervention. On one hand, it will help to elucidate if the response to an intervention in specific periods (e.g., in-school period) was replaced or compensated by a less level of physical activity subsequently (e.g., out-of-school period) [10,11]. On the other hand, it will inform future interventions to focus on time-based effects on physical activity in youth.

To fill in the research gaps, we conducted a nested study in the DECIDE-Children study [12] and aimed to examine whether a multifaceted intervention focusing on both children and contexts (schools and families) could improve the intensity and amounts of physical activity within specific periods of a week (i.e., weekday/weekend day) or a day (i.e., in-school/out-of-school periods) by using the objective accelerometer measure. We also examine whether potential moderators were associated with the effect size of the intervention.

## Methods

### The study context

The previous main study, the Diet, ExerCIse and CarDiovascular hEalth (DECIDE-Children) has been described in detail [12, 13] and prospectively registered at ClinicalTrials.gov (NCT03665857). Briefly, the DECIDE-Children study was a cluster-randomized controlled trial conducted from September 2018 to June 2019. A total of 24 primary schools from Beijing (8 schools), Changzhi of Shanxi province (8 schools), and Urumqi of Xinjiang province (8 schools) were recruited into the study and randomly allocated as 1:1 to the intervention (12 schools, 705 children) or control (12 schools, 687 children) arms. The intervention reduced body mass index (BMI) and other adiposity outcomes (e.g., BMI Z-score, body fat percentage, waist circumference, the prevalence of obesity), improved dietary behaviors and obesity-related knowledge, but it did not increase self-reported time spent on moderate-to-vigorous-intensity physical activity (MVPA) [13].

### The present study

The nested study was conducted as part of the DECIDE-Children study. To be eligible for the present study, children had to have consented to participate in the DECIDE-Children study and had their physical activity measured by an accelerometer. According to the pre-specified plan for data collection, accelerometer data were collected only from a subgroup nested in the DECIDE-Children study, i.e., 8 primary schools in Beijing, with 4 schools in the Dongcheng district (located in the center of the city) and 4 schools in the Mentougou district (located in a rural suburban area). One class was selected from Grade 4 in each school. Written informed consent was obtained from child-parent dyads. We obtained ethical approval from the Peking University Institution Review Board (IRB00001052-18021).

### Intervention arm

The multifaceted intervention was developed based on the Social-Ecological Model and has been reported previously [12, 13]. Briefly, the intervention included three child-oriented components to promote a healthy diet and physical activity (health education, reinforcement of physical activity, and BMI monitoring and feedback) and two context-oriented components to engage schools and families in facilitating children’s behavioral changes.

Two key messages were delivered to children and parents throughout the intervention:

- Children should do sufficient **amounts** of physical activity every day, at least 60 minutes/day;
- The **intensity** of physical activity should be moderate to vigorous.

Reinforcement of physical activity within the school was aimed at achieving the goal of “One-Hour Physical Activity on Campus Every School Day” as recommended by both the Chinese national government and WHO guideline [3]. Schools were required to ensure adequate time for three types of physical activity sessions during school hours, including physical education classes, class-break exercise, and extracurricular activities. Physical education teachers were trained by the project staff to deliver physical activity sessions achieving moderate to vigorous intensity. To encourage children to be physically active outside school, comic books were disseminated to children and parents which demonstrated various types of physical activity (e.g., strength training, jumping, running, and exercise games) using cartoon characters. Children could also do exercise by following videos periodically updated in the smartphone application.

### Quality control and process evaluation of the physical activity components of the intervention

Trained project staff went to each intervention school weekly to observe and record the amounts and intensity of in-school physical activity sessions, according to ‘An Operation Manual for Project Staff Involved in the Intervention’ [12]. Based on these field observations, the intervention schools achieved the amounts of 60 minutes/day of physical activity for a median (range) of 83% (62 to 97%) of school days, and achieved the intensity of MVPA in 88%, 84%, and 58% of physical education classes, extracurricular activities, and class-break exercises [13]. Based on parental reports of their involvement in reinforcing children’s physical activity outside school, 2.0%, 4.4%, 27.3%, 49.3%, and 17.1% of parents “never”, “very little”, “sometimes”, “often”, and “always” reinforced children’s physical activity outside school, respectively [13].

### Control arm

Schools in the control arm continued their usual health education sessions without activities focusing on childhood obesity prevention.

### Collection and processing of accelerometer data

Baseline and follow-up assessments of physical activity were undertaken in September 2018 (before randomization) and June 2019 (end of the trial) respectively. Each child participating in this nested study received an accelerometer (Actigraph GT3XP) and instructions on appropriate use. Children were shown by trained project staff to properly wear the device around their waist and were asked to wear the device for 7 consecutive days except during water-contact activities (e.g., bathing, swimming) and sleep. The Actigraph captured 3-dimensional acceleration every 15 seconds. The ActiLife (V6.8.2) software was used to assist with processing of accelerometer data.

### Physical activity outcomes

Study outcomes included intensity and amounts of physical activity at the end of the trial. The intensity of physical activity was classified using accelerometer count thresholds developed by Evenson et al [14]. Accelerometer counts were translated into minutes of sedentary, light-, moderate-, and vigorous-intensity physical activity using thresholds of 0-100 counts/minute, 101-2295 counts/minute, 2296-4011 counts/minute, and >4011 counts/minute, respectively [14]. The amounts of physical activity were calculated by summing the minutes of light-, moderate-, and vigorous-intensity physical activity for each day. All outcomes were assessed for weekdays or weekend days separately due to possibly distinct activity patterns [15]. Weekday physical activity outcomes were further analyzed specific to 3 time segments: within the waking hours of the whole day, in-school hours (8:00 AM∼4:59 PM), and out-of-school hours (5:00 PM∼11:00 PM).

### Moderators

Based on systematic reviews [7, 16], we conducted 7 pre-specified subgroup analyses to assess whether intervention effects on the physical activity outcomes differed by sex (boy, girl), age (≥ 9.6 years, < 9.6 years; based on the median age), baseline BMI status (overweight/obese; not overweight/obese), baseline in-school time spent on MVPA (≥ 3.01 min/hour, < 3.01 min/hour; based on the median value), as well as accelerometer compliance represented by valid wearing days during weekdays (6 days, < 6 days; based on the median value) and wearing time per weekday (≥ 819.23 min/day, < 819.23 min/day; based on the median value).

### Other measures

Height and body weight data were collected by physical examination before randomization. BMI (kg/m^2^) was then calculated as body weight (kg) divided by the square of height (m). Overweight and obesity were defined according to both Chinese national screening criteria (i.e., age- and sex-specific BMI cut-offs) [17] and the WHO growth reference (i.e., overweight: BMI Z-score > 1; obesity: BMI Z-score > 2) [18]. Demographic data (e.g., maternal education) were also collected before randomization from the parent questionnaire.

### Statistical analyses

Baseline characteristics at the cluster (school) and individual levels were described as means [standard deviations (SDs)] or medians (range) for continuous variables, and frequencies (percentages) for categorical variables. Linear mixed regression models allowing for the school-clustering effect (i.e., schools treated as random factors in the models) were used to estimate the effect size of the intervention. We built two models for each outcome. Model 1 (“the basic model”) was only adjusted for the baseline value of the outcome (e.g., daily time spent on the sedentary activity at baseline). Model 2 (“the adjusted model”) was additionally adjusted for children’s age, sex, and BMI at baseline.

We also conducted three sensitivity analyses by using different criteria for processing accelerometer data, as there has not been a unified criterion in terms of the definition of non-wear time within a day, a valid day, and a minimum number of valid wearing days per week [19, 20]. In main analyses, the non-wear time within a day was defined as more than 20 minutes of 0 count data, a valid day was defined as no less than 8 wearing hours (480 minutes), and the minimum number of valid wearing days was defined as 3 weekdays and 1 weekend day to represent ‘usual’ activity. In sensitivity analyses, non-wear time was re-defined as 60 minutes of 0 count data (sensitivity analyses 1), a valid day was re-defined as no less than 7 wearing hours (420 minutes) (sensitivity analyses 2), and the minimum number of valid wearing days was re-defined as 2 weekdays and 1 weekend day (sensitivity analyses 3).

Additionally, we conducted 7 pre-specified subgroup analyses to assess whether the intervention effects differed by the subgroup variables (i.e., moderators, described in “Moderators”). We used interaction terms between each subgroup variable (i.e., moderator) and group assignment (intervention or control) to examine the possibly differential intervention effects according to subgroups. Statistical significance was defined as a 2-sided *P* value of less than 0.05. R software (version: 4.1.3; http://www.r-project.org/) was used for the statistical analyses.

## Results

In this nested study, 218 consenting children (119 in intervention and 99 in control) for the 8 schools were selected from the main study, 154 (70.6%) completed assessments of physical activity and had valid accelerometer data at baseline, and 98 (63.6%) remained in the study and had valid accelerometer-assessed physical activity data at the end of the trial (Figure 1). Characteristics of the study participants (n = 98) included in this study did not differ significantly from those excluded (n = 56) (Supplemental Table 1). Most baseline characteristics of selected schools and children were similar between the intervention and control arms, except that children in the intervention arm were more likely to be in the Dongcheng district (56.7%) than the control children (41.9%) (Table 1).

**Figure 1.**
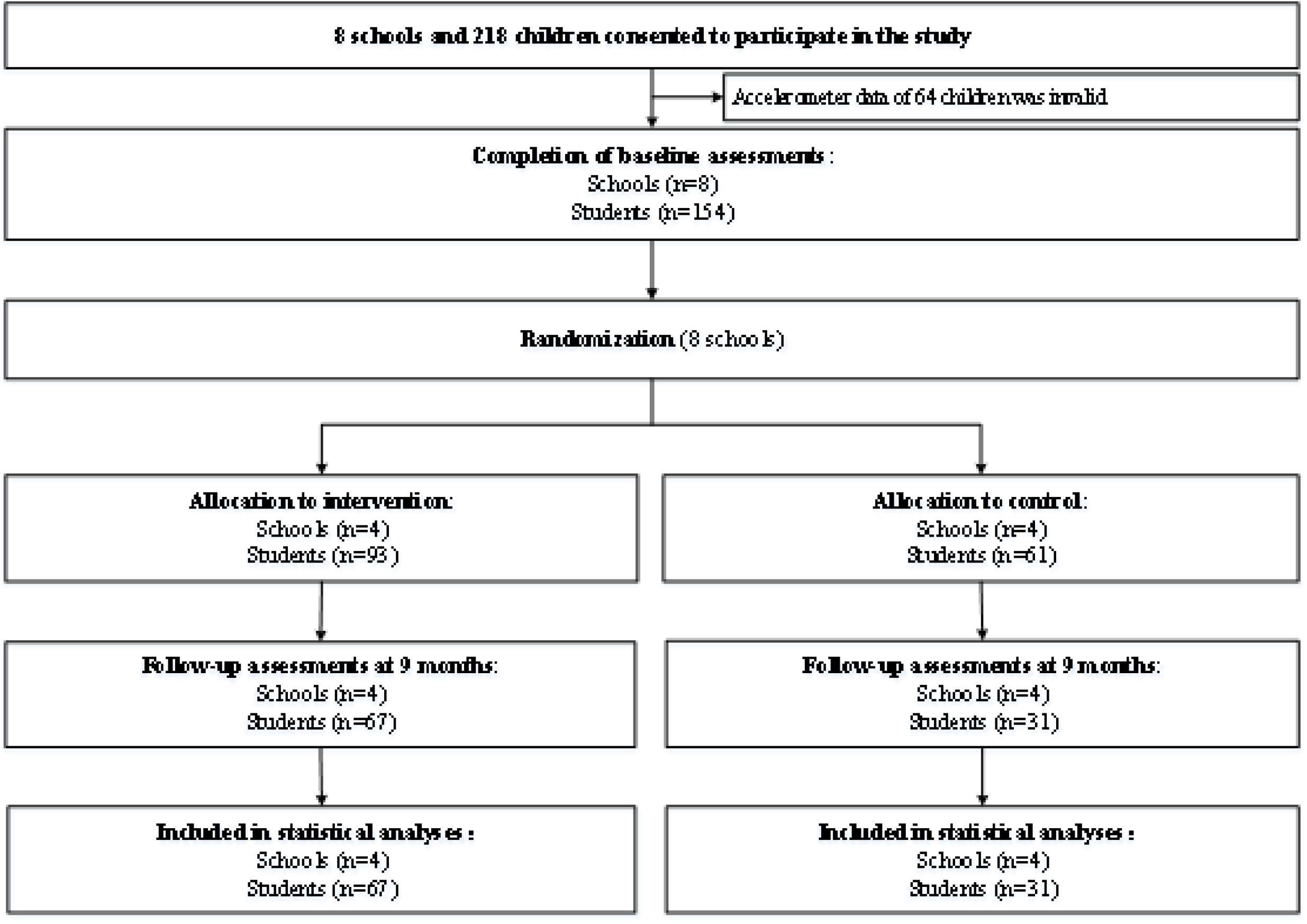
Study flowchart of the nested study.

**Table 1.**
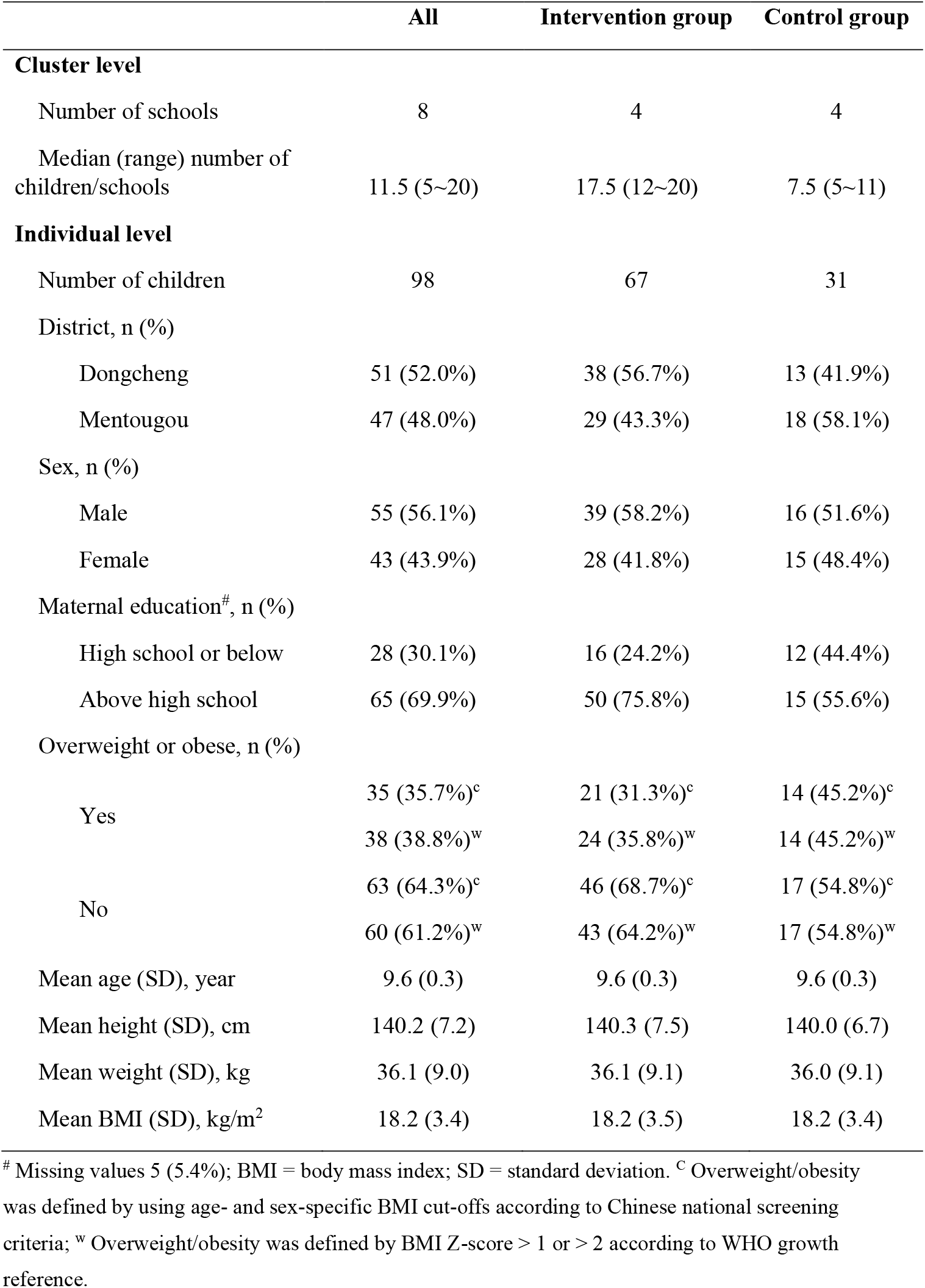
Baseline characteristics of schools and children, overall and by trial arm.

|  | All | Intervention group | Control group |
| --- | --- | --- | --- |
| **Cluster level** |  |  |  |
| Number of schools | 8 | 4 | 4 |
| Median (range) number of children/schools | 11.5 (5~20) | 17.5 (12~20) | 7.5 (5~11) |
| **Individual level** |  |  |  |
| Number of children | 98 | 67 | 31 |
| District, n (%) |  |  |  |
| Dongcheng | 51 (52.0%) | 38 (56.7%) | 13 (41.9%) |
| Mentougou | 47 (48.0%) | 29 (43.3%) | 18 (58.1%) |
| Sex, n (%) |  |  |  |
| Male | 55 (56.1%) | 39 (58.2%) | 16 (51.6%) |
| Female | 43 (43.9%) | 28 (41.8%) | 15 (48.4%) |
| Maternal education<sup>#</sup>, n (%) |  |  |  |
| High school or below | 28 (30.1%) | 16 (24.2%) | 12 (44.4%) |
| Above high school | 65 (69.9%) | 50 (75.8%) | 15 (55.6%) |
| Overweight or obese, n (%) |  |  |  |
| Yes | 35 (35.7%)<sup>c</sup> | 21 (31.3%)<sup>c</sup> | 14 (45.2%)<sup>c</sup> |
|  | 38 (38.8%)<sup>w</sup> | 24 (35.8%)<sup>w</sup> | 14 (45.2%)<sup>w</sup> |
| No | 63 (64.3%)<sup>c</sup> | 46 (68.7%)<sup>c</sup> | 17 (54.8%)<sup>c</sup> |
|  | 60 (61.2%)<sup>w</sup> | 43 (64.2%)<sup>w</sup> | 17 (54.8%)<sup>w</sup> |
| Mean age (SD), year | 9.6 (0.3) | 9.6 (0.3) | 9.6 (0.3) |
| Mean height (SD), cm | 140.2 (7.2) | 140.3 (7.5) | 140.0 (6.7) |
| Mean weight (SD), kg | 36.1 (9.0) | 36.1 (9.1) | 36.0 (9.1) |
| Mean BMI (SD), kg/m<sup>2</sup> | 18.2 (3.4) | 18.2 (3.5) | 18.2 (3.4) |

<sup>#</sup> Missing values 5 (5.4%); BMI = body mass index; SD = standard deviation. <sup>c</sup> Overweight/obesity was defined by using age- and sex-specific BMI cut-offs according to Chinese national screening criteria; <sup>w</sup> Overweight/obesity was defined by BMI Z-score > 1 or > 2 according to WHO growth reference.

Children’s physical activity patterns (assessed by accelerometer counts per minute) throughout the day differed between weekdays and weekend days. Figures 2 (A) and 2 (B) show weekday-specific patterns of physical activity at 10-minute intervals during waking hours, which were characterized by peaks and troughs throughout school hours. In comparison, the weekend-specific patterns of physical activity shown in Figure 2 (C) and (D) had far fewer fluctuations in terms of both frequency and amplitude throughout the day. Moreover, Figure 2 shows that weekday-specific patterns of physical activity within the “in-school period” differ from those within the “out-of-school period”.

**Figure 2.**
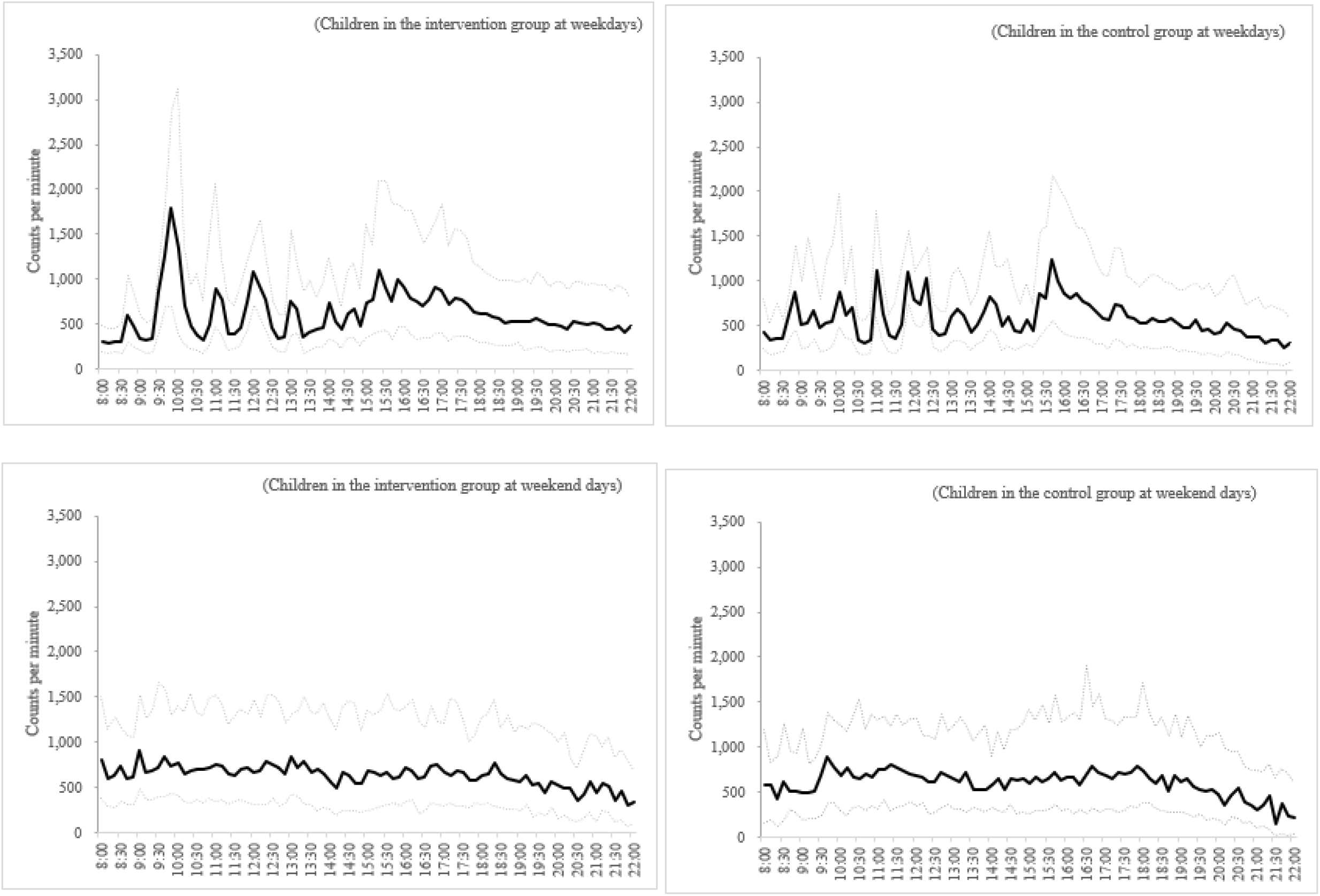
Weekday- and weekend-specific amounts of physical activity at baseline. (Solid line: the median values of counts per minute; Dotted line: the 1^st^ and 3^rd^ quartile values of counts per minute.)

### Intervention effects on weekday-specific physical activity

The study found that children in the intervention group spent more time on MVPA within the in-school period than those in the control group (Table 1; adjusted mean difference: 0.54 minutes/hour, 95% CI: 0.13, 0.94, *P* = 0.012). However, no significant differences were found in the weekday-specific physical activity outcomes within the whole day between the intervention and control arms (*P* > 0.05). The two statistical models showed similar results of the intervention effects on weekday-specific physical activity. Results from sensitivity analyses by using varied criteria for accelerometer data processing remained similar to those from main analyses (Supplemental Table 2∼4).

As shown in Figure 3, the effect size of weekday-specific MVPA within the in-school period did not differ significantly by sex, baseline BMI status, baseline MVPA, or accelerometer compliance. However, the effect size was stronger among children who were younger than 9.6 years than the older (adjusted mean difference: 0.83 [95% CI: 0.26, 1.40] vs 0.09 [95% CI: -0.46, 0.64]; *P* = 0.035 for interaction).

**Figure 3.**
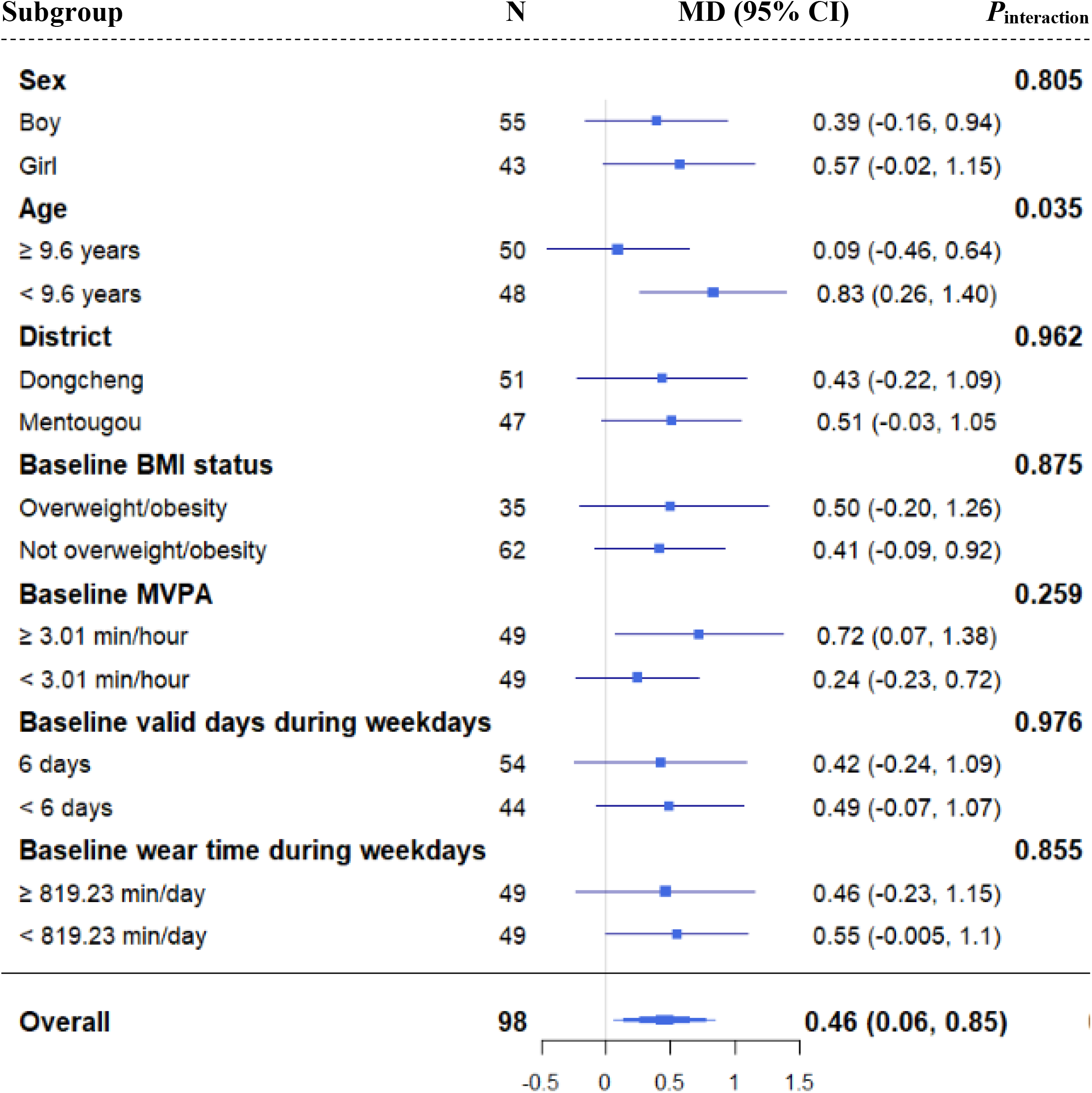
Subgroup analyses of intervention effects on the weekday-specific MVPA within the in-school period.

### Intervention effects on weekend-specific physical activity

Table 3 shows the effects of the intervention on weekend-specific physical activity. No significant differences were found between the two arms on the daily time spent on the sedentary activity, light-intensity physical activity, MVPA, or the total amounts of physical activity during weekend days at the end of the trial (all *P* > 0.05). Results from sensitivity analyses were similar to those from main analyses (Supplemental Table 2∼4).

**Table 2.** Intervention effects on weekday-specific physical activity, for the whole day, and during or out of school hours.

| Outcomes* | Intervention, mean (SD) |  | Control, mean (SD) |  | Intervention versus control |  |
| --- | --- | --- | --- | --- | --- | --- |
|  | Baseline | End of trial | Baseline | End of trial | Adjusted mean difference (95% CI) | P value |
| Within the whole weekday |  |  |  |  |  |  |
| Sedentary, minutes/day | 563.2 (65.3) | 539.0 (77.7) | 552.7 (71.9) | 547.5 (96.9) | M1: -16.40 (-49.68, 15.17) | 0.422 |
|  |  |  |  |  | M2: -19.78 (-61.51, 20.50) | 0.438 |
| Light PA, minutes/day | 211.3 (47.6) | 231.4 (46.9) | 216.9 (45.6) | 225.0 (69.1) | M1: 10.71 (-22.17, 44.30) | 0.585 |
|  |  |  |  |  | M2: 11.30 (-26.01, 49.37) | 0.602 |
| MVPA, minutes/day | 43.9 (18.7) | 47.5 (18.4) | 40.3 (12.1) | 40.2 (15.9) | M1: 4.98 (-0.94, 10.89) | 0.104 |
|  |  |  |  |  | M2: 4.53 (-1.02, 10.08) | 0.121 |
| Total PA, minutes/day | 255.2 (56.5) | 279.0 (55.9) | 257.2 (51.0) | 265.2 (80.6) | M1: 15.37 (-10.49, 39.79) | 0.449 |
|  |  |  |  |  | M2: 15.88 (-21.89, 54.54) | 0.493 |
| Counts per minute | 877.8 (206.8) | 927.9 (238.1) | 872.9 (190.3) | 878.5 (242.4) | M1: 44.78 (-26.17, 115.14) | 0.255 |
|  |  |  |  |  | M2: 40.57 (-26.22, 107.35) | 0.279 |
| Within school hours (8:00 AM~16:59 PM) |  |  |  |  |  |  |
| Sedentary, minutes/hour | 39.3 (5.6) | 34.9 (7.6) | 35.6 (7.3) | 32.7 (9.3) | M1: -0.16 (-3.39, 3.05) | 0.929 |
|  |  |  |  |  | M2: -0.03 (-3.1, 3.04) | 0.985 |
| Light PA, minutes/hour | 14.2 (3.9) | 14.9 (4.5) | 13.7 (3.7) | 13.5 (5.6) | M1: 1.19 (-1.31, 3.73) | 0.425 |
|  |  |  |  |  | M2: 1.27 (-1.51, 4.11) | 0.441 |
| MVPA, minutes/hour | 3.1 (1.2) | 3.0 (1.2) | 2.8 (1.1) | 2.3 (1.1) | M1: 0.54 (0.13, 0.94) | 0.012 |
|  |  |  |  |  | M2: 0.47 (0.08, 0.86) | 0.024 |
| Total PA, minutes/hour | 17.3 (4.5) | 17.9 (5.1) | 16.4 (4.3) | 15.8 (6.3) | M1: 1.73 (-0.82, 4.31) | 0.268 |
|  |  |  |  |  | M2: 1.76 (-1.05, 4.63) | 0.311 |
| Counts per minute | 863.5 (198.7) | 836.9 (220.5) | 808.5 (243.5) | 725.2 (251.1) | M1: 81.99 (-15.71, 179.21) | 0.144 |
|  |  |  |  |  | M2: 68.51 (-15.75, 152.83) | 0.179 |

| Out of school hours (17:00 PM~23:00 PM) |  |  |  |  |  |  |
| --- | --- | --- | --- | --- | --- | --- |
| Sedentary, minutes/hour | 24.6 (5.9) | 22.8 (7.6) | 22.4 (6.0) | 20.9 (8.0) | M1: 0.44 (-2.46, 3.35) | 0.766 |
|  |  |  |  |  | M2: 0.53 (-2.52, 3.6) | 0.756 |
| Light PA, minutes/hour | 9.6 (3.1) | 9.3 (3.5) | 8.4 (2.7) | 8.1 (3.5) | M1: 1.08 (-1.73, 3.92) | 0.497 |
|  |  |  |  |  | M2: 1.06 (-1.75, 3.91) | 0.504 |
| MVPA, minutes/hour | 1.7 (1.3) | 1.5 (1.0) | 1.1 (0.9) | 1.4 (0.9) | M1: -0.02 (-0.50, 0.46) | 0.931 |
|  |  |  |  |  | M2: -0.04 (-0.54, 0.46) | 0.875 |
| Total PA, minutes/hour | 11.2 (3.9) | 10.8 (4.1) | 9.6 (3.2) | 9.4 (4.2) | M1: 1.07 (-2.21, 4.39) | 0.562 |
|  |  |  |  |  | M2: 1.02 (-2.29, 4.37) | 0.584 |
| Counts per minute | 628.7 (242.3) | 575.4 (220.8) | 499.1 (181.6) | 504.0 (212.0) | M1: 10.86 (-117.44, 140.77) | 0.880 |
|  |  |  |  |  | M2: 10.46 (-114.68, 137.5) | 0.882 |

\*Sedentary, light, moderate, vigorous PA: defined as $\leq 100$ , 101-2295, 2296-4011, and $>4011$ accelerometer counts per minute, respectively. M1 refers to the basic model of linear mixed regression allowing for the school-clustering effect, with adjustment for baseline value of the outcome. M2 refers to the adjusted model of linear mixed regression on the basis of M1, with further adjustment for children’s age, sex, and BMI at baseline. Abbreviations: CI = confidence interval; MVPA = moderate-to-vigorous-intensity physical activity; PA = physical activity.

**Table 3.** Intervention effects on weekend-specific physical activity.

| Outcomes* | Intervention, mean (SD) |  | Control, mean (SD) |  | Intervention versus control |  |
| --- | --- | --- | --- | --- | --- | --- |
|  | Baseline | End of trial | Baseline | End of trial | Adjusted mean difference (95% CI) | *P* value |
| Sedentary, minutes/day | 487.6 (100.5) | 480.1 (106.1) | 472.1 (84.2) | 468.0 (123.0) | M1: 5.94 (-38.86, 50.74)<br>M2: 10.79 (-33.66, 55.24) | 0.797<br>0.643 |
| Light PA, minutes/day | 223.4 (66.4) | 226.9 (64.6) | 207.2 (48.5) | 201.7 (65.9) | M1: 18.48 (-7.19, 44.14)<br>M2: 19.29 (-4.79, 44.74) | 0.164<br>0.131 |
| MVPA, minutes/day | 41.6 (28.8) | 37.9 (25.6) | 37.4 (19.6) | 38.3 (28.7) | M1: -0.83 (-15.22, 13.77)<br>M2: -1.06 (-16.07, 14.17) | 0.915<br>0.897 |
| Total PA, minutes/day | 265.0 (82.9) | 264.8 (79.7) | 244.6 (59.3) | 240.0 (85.8) | M1: 15.89 (-16.08, 48.31)<br>M2: 18.97 (-12.71, 46.44) | 0.335<br>0.405 |
| Counts per minute | 931.9 (324.6) | 917.5 (338.8) | 886.5 (264.2) | 907.1 (382.3) | M1: -7.47 (-164.7, 150.98)<br>M2: -16.19 (-178.35, 147.06) | 0.929<br>0.854 |

\*Sedentary, light, moderate, vigorous PA: defined as ≤100, 101-2295, 2296-4011, and >4011 accelerometer counts per minute, respectively. M1 refers to the basic model of linear mixed regression allowing for the school-clustering effect, with adjustment for the baseline value of the outcome. M2 refers to the adjusted model of linear mixed regression on the basis of M1, with further adjustment for children's age, sex, and BMI at baseline. Abbreviations: CI = confidence interval; MVPA = moderate-to-vigorous-intensity physical activity; PA = physical activity.

## Discussion

The present study, nested in a rigorously designed cluster-randomized controlled trial, found that the intervention was effective in improving the in-school physical activity during weekdays, and the effect size did not differ by sex, baseline BMI status, baseline MVPA, or accelerometer compliance. But we observed no intervention effects on weekend-specific or out-of-school physical activity outcomes. Our findings were echoed by the results of the KISS study [21], in which the intervention was beneficial on facilitating in-school MVPA [1.19 physical activity z-score (95% CI: 0.78, 1.60)] but not on out-of-school MVPA [-0.06 (95% CI: -0.39, 0.27) physical activity z-score].

Our findings of intervention effects on time-specific physical activity could be interpreted by “the activitystat hypothesis” [22], that the more physical activity of children at one time seems to be compensated with less activity at another. Alternatively, children were more likely to be responsive to the structured sessions of physical activity organized by the school settings (e.g., physical education classes, extracurricular activities, and class-break exercise) than the non-compulsory intervention elements outside school (e.g., family homework) [23]. Another interpretation of the finding lies in the family members who influenced much on children’s physical activity behavior, yet it is not a simple task to sufficiently target behavioral change for the whole family [10].

Of note, the effect size of intervention effects in in-school MVPA was greater among subgroups of children who were younger. A previous meta-analysis also found that younger children benefited more from the physical activity intervention [16]. Studies from cross-sectional and prospective observations indicated that physical activity declined with age [24]. These consistent findings provided important insights that future intervention strategies should be tailored to children with age differences.

Despite the intriguing findings of physical activity within specific periods, we found no differences in the whole day accelerometer-assessed MVPA between the intervention and control groups. This finding was consistent with the main study finding that relied on questionnaire-based physical activity measure [13]. Our findings were also mirrored by two meta-analysis studies [6,7] and the subsequent rigorously designed trials undertaking a school-based approach to tackle childhood obesity similar to ours [25-27]. Several lines of evidence indicated that changing the whole physical inactivity patterns by using the current intervention approach in primary school children is far from satisfactory.

### Strengths and limitations

Childhood is the critical life stage for establishing healthy behaviors such as physical activity. Motivating children to be physically active was just a beginning point for this DECIDE-Children intervention which lasted for only one school year. Future longer-term interventions should gradually increase the intensity of physical activity to maximize the health benefits. This nested study was limited by the small sample size due to practical barrier of collecting accelerometer data. However, our findings were consistent with previous meta-analysis studies with larger sample sizes [6, 7]. And the consistency in the results from several sensitivity analyses made our study findings less likely to be biased. Additionally, the accelerometer, as the objective measurement tool of physical activity, cannot measure water-based (e.g., swimming) or cycling activities, but this weakness may have little influence on the validity of the findings since the physical activity components in the intervention did not specifically focus on increasing water-based or cycling activities [12]. Compliance is a common issue in studies assessing accelerometer-based physical activity [16]. The follow-up investigation of our study is not exceptional with the attrition rate of 36.4%. But characteristics of the sample were similar between those in the study and those excluded from the study.

Our study was unique in its data collection nested in a rigorously designed cluster-randomized controlled trial, physical activity assessed objectively by the accelerometer, and physical activity observed specifically on weekdays/weekend days as well as during in-school/out-of-school hours. The time-segment-specific findings from this study can inform the future design of a school-based intervention. It might be an ideal option to pay equal attention to out-of-school period in addition to in-school period, to lessen the compensatory issue of less physical activity during the out-of-school time.

## Conclusions

The childhood obesity intervention effectively increased MVPA during in-school hours, and younger children appeared to benefit more from the intervention. Future interventions delivered via the school settings should focus more on children’s physical activity outside school time and tailor intervention strategies to children’s age difference, in order to maximize the intervention effects on improving the whole patten of children’s physical activity.

## Data Availability

All data produced in the present study are available upon reasonable request to the authors.

## Abbreviations

BMI: body mass index
CI: confidence interval
DECIDE: the Diet, ExerCIse and CarDiovascular hEalth
IQR: inter-quartile range
MVPA: moderate-to-vigorous-intensity physical activity
OR: odds ratio
SD: standard deviation
WHO: world health organization

## Declarations

### Authors’ contributions

*Study design*: HJW and ZL; *Data collection*: ZL, ZHY, SZ, AYG, and FZ; *Quality assessment*: ZL and HJW; *Data analysis*: ZL, ZHY, JZ, and HJW; *Drafting of the manuscript*: ZL, ZHY, LMW, JZ, SZ, AYG, FZ, and HJW; *Critical revision of the manuscript for important intellectual content*: ZL, LMW, and HJW.

## Acknowledgments

We thank all children, parents, and school teachers for their participation in the program, and the study team for the DECIDE-children study.

---

**Supplemental Table 1 Comparison of characteristics between the remaining children and those without follow-up assessment of weekend-specific physical activity in the nested study**

**Supplemental Table 2 Intervention effects on weekday- and weekend-physical activity (sensitivity analyses 1)**

**Supplemental Table 3 Intervention effects on weekday- and weekend-physical activity (sensitivity analyses 2)**

**Supplemental Table 4 Intervention effects on weekday- and weekend-physical activity (sensitivity analyses 3)**

